# PPIE in clinical trials at scale: analysis of the first 3,250 responses on the POrtal for Patient and Public Engagement in Dementia (POPPED)

**DOI:** 10.1101/2025.08.18.25333896

**Authors:** Hongyi Qin, Linda Pointon, James Carpenter, Vanessa Raymont, Ross Dunne, Suzanne Reeves, Shabinah Ali, Sabahat Iqbal, Cristina Bonet-Olivares, Jo Whittle, Laura Rizzo, Paresh Malhotra, Benjamin R Underwood, AD-SMART team

**Author notes:** Joint corresponding authors. Editorial office correspondence: Benjamin R Underwood, Department of Psychiatry, University of Cambridge, Cambridge, CB2 0SZ, UK.

## Abstract

**INTRODUCTION:** Patient and public involvement and engagement (PPIE) is essential for improving research but is often limited in scale. This study explored the potential for large-scale involvement using a web-based approach.

**METHODS:** We created an online portal to collect feedback on dementia research and sought views on the forthcoming UK-based adaptive platform trial testing repurposed drugs for Alzheimer’s disease (AD-SMART). Participants completed a survey with a ranking task for four anonymized drugs prioritised for inclusion and discrete choice experiments (DCEs) on treatment attributes trade-offs.

**RESULTS:** Responses from over 3,250 people across 27 countries and 6 continents strongly supported for the trial. *Metformin* was the most preferred, followed by *Atomoxetine*, *Isosorbide Mononitrate*, and *Levetiracetam*. Safety ranked highest, followed by evidence of efficacy and convenience. Analyses were stratified by sex, age, and dementia experience.

**DISCUSSION:** Web-based PPIE can effectively inform dementia research at scale and offers a transferable model for other studies.

## 1. Background

Patient and public involvement and engagement in research (PPIE) is a process by which patients and the public are active partners in all stage of the research cycle, from setting priorities and research questions, to design and dissemination of findings.^1,2^ Standards exist for PPIE based on principles of inclusivity, support, working together, governance, communication and impact.^3,4^ Further guidance exists on measuring the impact of PPIE and publishing findings.^5,6^

Despite this progress, there is still considerable room for development.^7^ First, many PPIE groups are often small in number and may not include people from underserved groups.^8^ This limitation is particularly concerning in dementia research, where an estimated 60 million people worldwide living with the condition and increasing numbers of families and caregivers are affected.^9^ In some cases, PPIE is limited, focusing on dissemination rather than involving collaborators in a more integrated and participatory role.^10,11^ More meaningful engagement would include working with PPIE contributors throughout the research process from initial designing to implementation.^12^ In other areas of medicine, researchers have attempted to address these limitations by engaging online with a global community.^13^ However, whether web-based approaches can achieve scalable, inclusive PPIE in dementia research remains unknown. To address this gap, we investigated the potential for engaging with patients and the public at scale using a novel purpose-built web-based portal to facilitate mass feedback.

This study was part of a wider initiative to design and implement a UK-wide platform in clinical Alzheimer’s Disease (Alzheimer’s Disease-Systematic Multi-Arm Adaptive Randomised Trial, (AD-SMART)) designed in early 2025. A key consideration for AD-SMART is which candidate drugs should be prioritised for initial evaluation in the platform. Within this broader context, our specific objectives were to (1) explore patient and public preferences regarding the initial evaluation of four anonymised, real-world candidate drugs included in the AD-SMART trial; (2) examine how individuals trade off between key medications attributes, such as side effects and dosage regimen, and to (3) assess whether and how these preferences vary across subgroups with different demographic backgrounds.

Given the scale of AD-SMART (approximately 400 patients per arm) and the critical importance of patient and public involvement, this represented an ideal opportunity to pilot and evaluate a novel web-based approach to determine whether it would be feasible to obtain involvement and engagement at scale.

## 2. Methods

### 2.1. Study settings

The broader context for this study is the AD-SMART trial, an adaptive platform trial designed to evaluate multiple treatments for a single disease with the ability to add or remove treatments being tested over time dependent on interim results.^14^ Members of the wider team have successfully delivered such platforms in oncology and neurodegenerative diseases.^15–17^ The development of a systematic, sustainable drug prioritisation pipeline is described in detail elsewhere.^18^ Briefly, this process involved engaging the wider AD community and inviting proposals for candidate treatments using a pre-specified template, followed by expert panel meetings. Artificial Intelligence (AI) guided reviews of the available literature were then used to create extended drug profiles for shortlisted drugs before further expert panel reviews with individualised rankings. Alongside this, a PPIE group provided qualitative feedback on the shortlisted drugs, based on lay summaries produced by the drug prioritisation team.

### 2.2. PPIE activities

PPIE was integrated throughout the preparatory stages of the online portal. Prior to the launch of the web-based survey, a series of PPIE meetings were conducted with a group of 14 individuals with lived experience of dementia, who had previously been involved in the AD-SMART drug prioritisation process. These contributors helped co-develop key elements of the study, including the structure and language of the web-based survey and the design of the online portal. They advised on the clarity, user-friendly language, and accessibility of the materials, with particular attention to how uncertainty and risk were communicated. Participants also provided feedback on the overall cognitive and emotional burden of completing the survey, leading to several adjustments to improve flow, comprehension, and usability. Importantly, people shared their views on each of the shortlisted drugs, including which they found more or less appealing and why. These in-depth qualitative insights helped inform the selection of attributes and levels used in the forthcoming survey, ensuring that the design was grounded in real-world perceptions and concerns expressed by people with lived experience.

In terms of engagement strategy, we used multiple channels to reach diverse audiences. This included continued collaboration with the original PPIE groups, outreach via social media platforms, translation in to Mandarin Chinese, and promotion in partnership with dementia-related charities and the Join Dementia Research platform. The engagement was inclusive of a broad range of voices, including people living with dementia, family carers, professionals working in dementia, and other members of the public with relevant perspectives. These efforts aimed to ensure the accessibility and relevance of the study, and to support a more representative model of PPIE in research.

### 2.3. Webpage and survey design

The POPPED portal (**PO**rtal for the **P**atient and **P**ublic **E**ngagement in **D**ementia Research, https://popped.org.uk/)^19^ was designed to host studies looking to obtain broader PPIE on dementia research. The study protocol was approved by the research ethics committee of the University of Cambridge (REC Reference: HBREC.2025.01). All data was administered using the REDCap platform and database securely hosted by the University of Cambridge.

Through the POPPED, we conducted a multi-language online survey in English and Mandarin Chinese, with additional languages planned for future development. The survey aimed to gain opinions on the four highest-ranked compounds shortlisted by the external panel for inclusion in AD-SMART, and apply this to future arms of the platform. Specifically, the survey consisted of three main sections: (1) socio-demographic background and their attitudes toward clinical trials that test more than one drug simultaneously against a placebo, i.e., enthusiasm for a platform trial design, (2) a ranking task assessing participants’ overall preferences for four anonymized, real-world Alzheimer’s drugs with potential efficacy, and (3) a series of discrete choice experiments (DCEs) to explore trade-offs among specific treatment attributes. The full survey is available at https://popped.org.uk/ad-smart-survey/.^20^ Data collection was carried out between March 7 and July 15, 2025 (see Supplementary Figure S1 for a detailed response over time).

Eligible participants for the web-based survey were individuals aged 18 years or older who were able to read either English or Mandarin Chinese. All participants were provided electronic consent prior to participation. Information to aid drug selection consisted of two main parts. First, in the ranking task, participants were presented with four anonymized drug cards representing the leading medications currently under consideration for inclusion in the trial, based on the process outlined in the AD-SMART setting. Based on the drug profiles, each card presented information on what the drug is currently used for, its proposed mechanism of action, the stage of clinical research or use, its delivery method, any special monitoring requirements, and its known side effects. Figure 1(a) provides an example of a drug card, and all drug cards are detailed in Supplementary Figure S2. Participants were asked to rank these four drugs according to their overall preferences to inform prioritization discussions for the trial.

**Figure 1.**
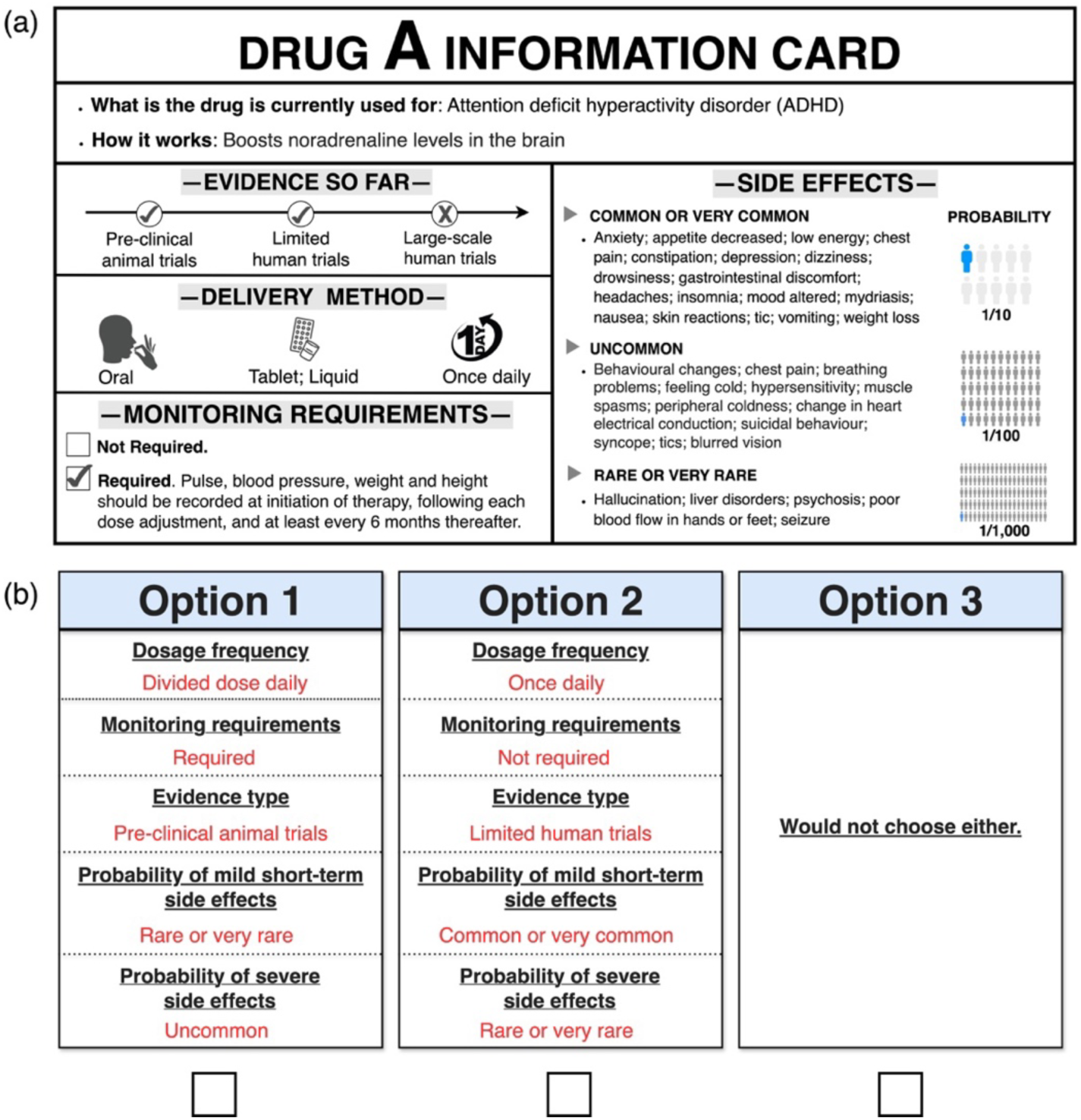
Materials used in the survey. (a). An example of the anonymized drug card. (b) An example of Choice task in DCEs.

Then, participants evaluated hypothetical drugs as opposed to actual treatments using a series of DCEs. These hypothetical profiles systematically varied across five key attributes identified in the preparatory PPIE sessions as being important: dosage frequency, monitoring requirements, type of supporting evidence, probability of mild short-term side effects, and probability of severe side effects. A detailed description of the attributes and levels is provided in Supplementary Table S1.

Based on all possible combinations of attribute levels, 72 pairwise choice scenarios could be constructed. We used a D-efficient experimental design to reduce respondents’ burden while maintain statistical efficiency. This type of design minimizes the determinant of the covariance matrix of the parameter estimates, thereby maximizing the information derived from the selected choice sets ^21^. Consequently, 12 tasks were required to achieve reliable estimation. To further minimize respondent fatigue, a block design was employed in which 12 tasks were divided into four subsets. Each participant was randomly assigned to complete three choice tasks plus one additional attention check, resulting in four tasks in total (see Supplementary Table S2). Figure 1(b) provides an example of a DCE choice task where participants chose between two hypothetical medications or selected neither option.

### 2.4. Statistical analysis

Full details on sample size calculation are provided in the Supplementary S.1. Based on our calculations, we set the total target sample size at approximately 2,800 participants to ensure sufficient power for subgroup analyses.

For the ranking task, raw ranking results were visualised using heatmaps to display the frequency of each drug being ranked in each position. Weighted preference scores were then calculated using a scoring scheme (rank 1 = 4 points, rank 2 = 3, rank 3 = 2, rank 4 = 1). The total weighted score for each drug was calculated as:

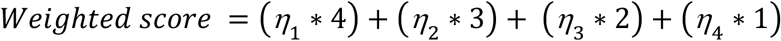

where η_1_ - η_4_ are the number of times the drug was ranked 1^st^ through 4^th^. Bar charts were used to present the total weighted scores.

For the DCE, a mixed logit model was used to quantify respondents’ preferences for medication attributes and to account for preference heterogeneity across individuals. The utility *U*_*i*j*t*_of respondent *i* from choosing medication *j* in choice set *t* is specified as:

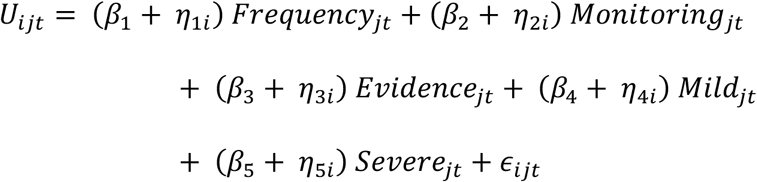

Where *Frequency*_j*t*_, *Monitoring*_j*t*_, *Evidence*_j*t*_, *Mild*_j*t*_, *Severe*_j*t*_ are dummy-coded attribute levels. *β*_1_ to *β*_5_ are the mean utility weights for attributes *k* relative to their reference levels, while *η*_*ki*_ capture the individual-specific random deviations. The *∈*_*i*j*t*_ is assumed to follow a Type I extreme value distribution.

Relative importance for each attribute was calculated based on the range of its estimated level coefficients, following the method described by Malhotra and Birks.^22^ The importance weight for attribute *k* was determined by:

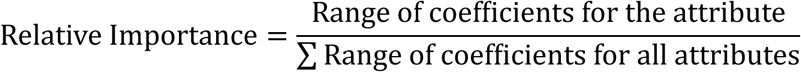

We then conducted two subgroup analyses to explore whether preferences for medication attributes differed across population subgroups. The first analysis stratified participants by age and gender, while the second analysis stratified them by dementia-related experience. For each subgroup, we estimated separate models and between-group comparisons. All analyses were conducted using Stata.

To examine whether the collected sample size was sufficient for stable parameter estimation, we also conducted a post hoc stability check by estimating the model with progressively larger subsamples (e.g., the first 500, 1000, 1500, and all respondents. See Supplementary S.2). The results suggested that the key parameter estimates stabilized after approximately 1000-1500 respondents, supporting the adequacy of the final sample.

## 3. Results

### 3.1. Sample characteristics

A total of 3,251 respondents were included in the analysis. Responses were received from 27 countries across 6 continents, with the top five being the United Kingdom, China, the United States, Ireland, and Australia. A world map showing countries with survey responses is available in Supplementary Figure S3.

As shown in Table 1, the largest age group was those aged 65-74 years, accounting for 31.28% of the sample. Overall, 71.2% of the total sample were aged 55 years and over, and 28.8% were aged 18-54 years. A majority identified as female (68.0%), while 31.3% identified as male. Most respondents indicated that their friends or family members had/have dementia (46.4%), followed by those who care for or have cared for someone with dementia (16.7%). Among those who reported having dementia, the majority (75.5%) were aged between 65 and 84 years, with 31.1% in the 65-74 age group and 44.4% in the 75-84 age group. Only a small proportion were younger than 55 years (10.0%) or aged 85 and over (3.3%).

**Table 1.** Participants characteristic.

| Characteristics |  | Frequency (Percentage)<br>N=3,251 |
| --- | --- | --- |
| **Age, years** |  |  |
|  | 18-24 | 175 (5.4%) |
|  | 25-34 | 283 (8.7%) |
|  | 35-44 | 171 (5.3%) |
|  | 45-54 | 306 (9.4%) |
|  | 55-64 | 808 (24.9%) |
|  | 65-74 | 1017 (31.3%) |
|  | 75-84 | 453 (13.9%) |
|  | 85 and over | 38 (1.2%) |
| **Gender** |  |  |
|  | Female | 2209 (68.0%) |
|  | Male | 1018 (31.3%) |
|  | Another gender | 14 (0.4%) |
|  | Prefer not to answer | 10 (0.3%) |
| **Participants' description** |  |  |
|  | My friend or family member had/has dementia | 1507 (46.4%) |
|  | I care/have cared for someone with dementia | 544 (16.7%) |

| **Characteristics** | **Frequency (Percentage)**<br>**N=3,251** |
| --- | --- |
| My work is related to dementia | 182 (5.6%) |
| I have dementia | 90 (2.8%) |
| None of the above | 883 (27.2%) |
| Prefer not to say | 45 (1.4%) |
| **Residence region** |  |
| United Kingdom | 2842 (87.4%) |
| China | 375 (11.5%) |
| United States | 5 (0.2%) |
| Ireland | 4 (0.1%) |
| Australia | 3 (0.1%) |
| Others | 22 (0.7%) |

Overall, our sample demonstrated broad demographic and experiential diversity, supporting the robustness and potential external relevance of the study findings across different age groups, genders, personal experiences with dementia, and geographic contexts.

### 3.2. Preferences ranking results

#### 3.2.1. Overall preferences

Figure 2 shows the distribution of participants’ preferences for each drug. The heatmap on the left shows that *Metformin* was most frequently selected as the top choice (1st rank: n=1,155, 35.5%), followed by *Isosorbide Mononitrate* (n=892, 27.4%), *Atomoxetine* (n=837, 25.7%), and *Levetiracetam* (n=367, 11.3%). *Levetiracetam* was most commonly chosen as the 4th preference (n=1,222, 37.6%), indicating a tendency for it to be ranked lower compared to other options. The bar chart on the right summarizes the weighted preference scores. The results indicate that *Metformin* accumulated the highest total weighted counts at the top rank (total weighted count=9,374), followed by *Atomoxetine*, *Isosorbide Mononitrate*, and *Levetiracetam*. Notably, even after weighting, *Metformin* remained predominantly favored as a first-line option, while *Levetiracetam* was more often considered suitable as a last choice.

**Figure 2.**
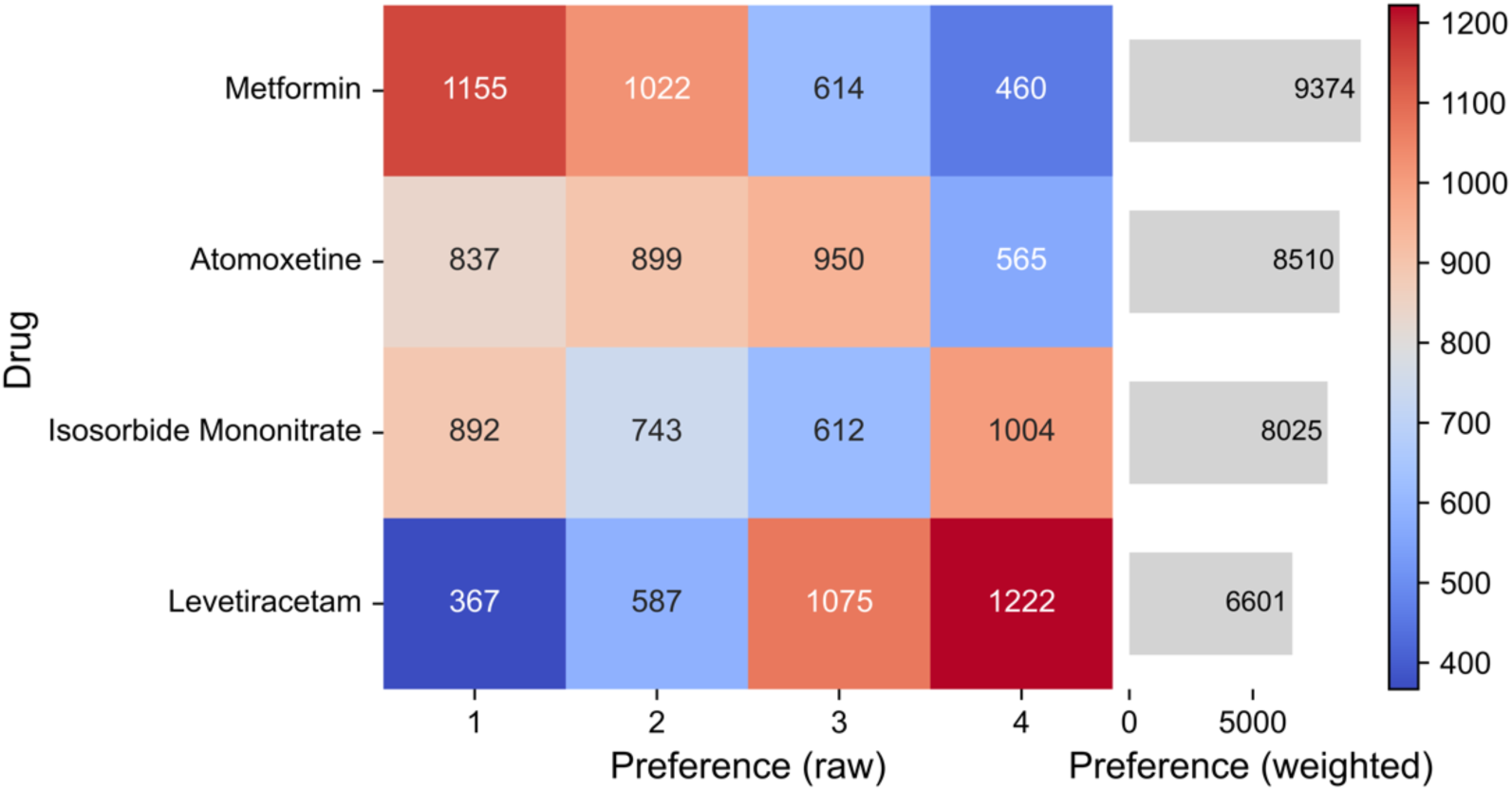
Preferences for four drugs across ranking positions. The heatmap (left) displays the raw counts of each drug ranked 1st to 4th by participants, while the bar chart (right) shows the total weighted preference scores for each drug.

#### 3.2.2. Subgroup preference ranking

##### Age and gender

To further explore potential demographic heterogeneity in drug preferences, we stratified the sample by age and gender. The threshold of 65 years was selected for two reasons. First, our data showed that the majority of respondents with dementia (78.8%) were aged over 65, indicating a concentration of samples in this age range. Second, 65 years old is a widely used clinical cutoff for young-onset dementia or working-age dementia.^23,24^ Figure 3 presents results for each subgroup.

**Figure 3.**
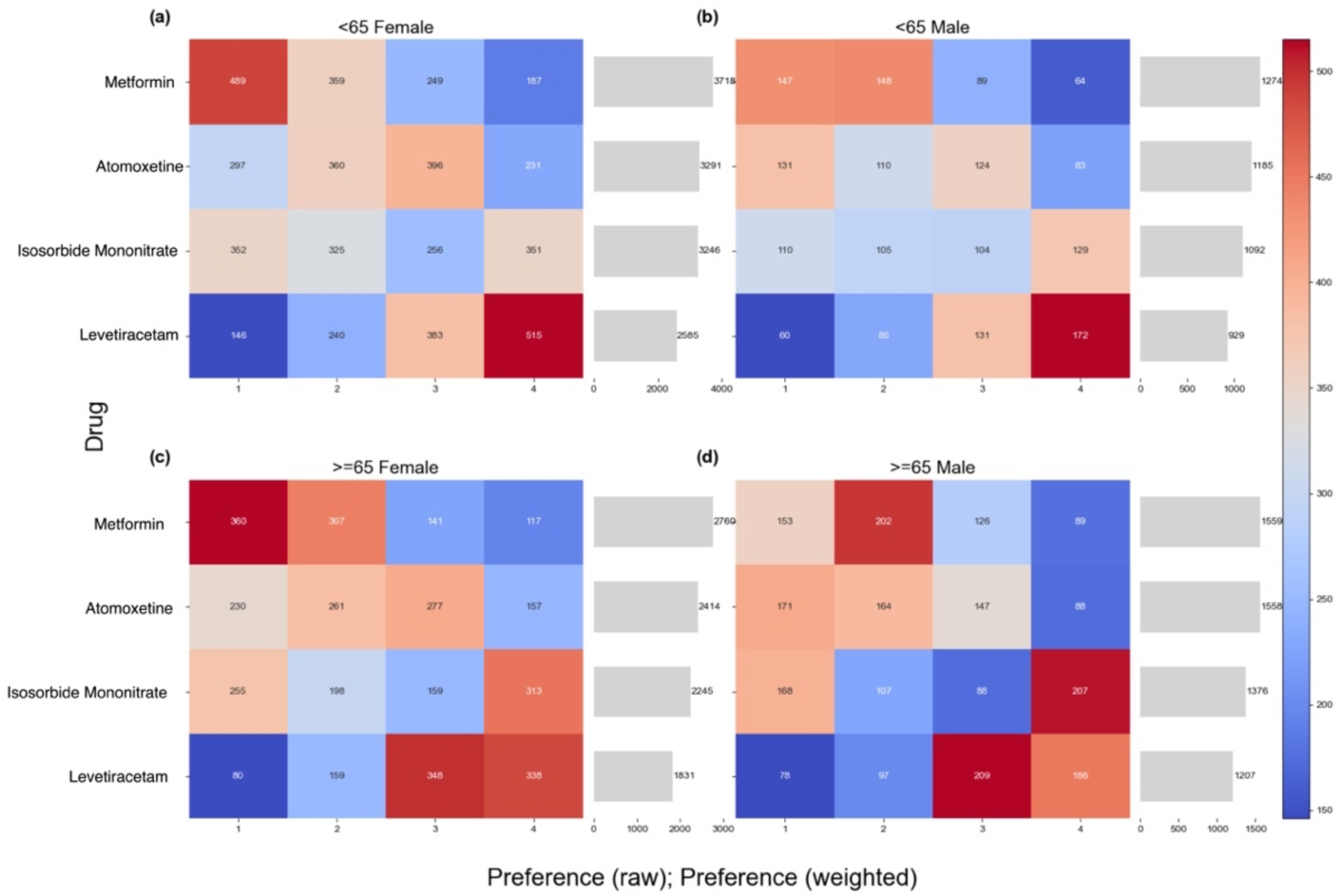
Preferences by age and gender. The heatmaps show the distribution of raw ranking frequencies for each drug. The bar charts show the corresponding total weighted preference scores. Results are presented separately for four participant groups: (a) <65 Female, (b) <65 Male, (c) ≥ 65 Female, and (d) ≥ 65 Male.

Across all four subgroups, the ranking pattern was consistent with that of the full sample: *Metformin* received the highest preference, followed by *Atomoxetine*, *Isosorbide Mononitrate*, and *Levetiracetam*. The weighted preference scores also suggest similar preference structures.

##### Dementia-related experience

In addition to demographic characteristics, we further explored whether people’s experience with dementia influences their preferences. Figure 4 presents rankings across participant subgroups based on their experience with dementia. Each panel displays raw preference distributions and the corresponding weighted scores for the four drugs.

**Figure 4.**
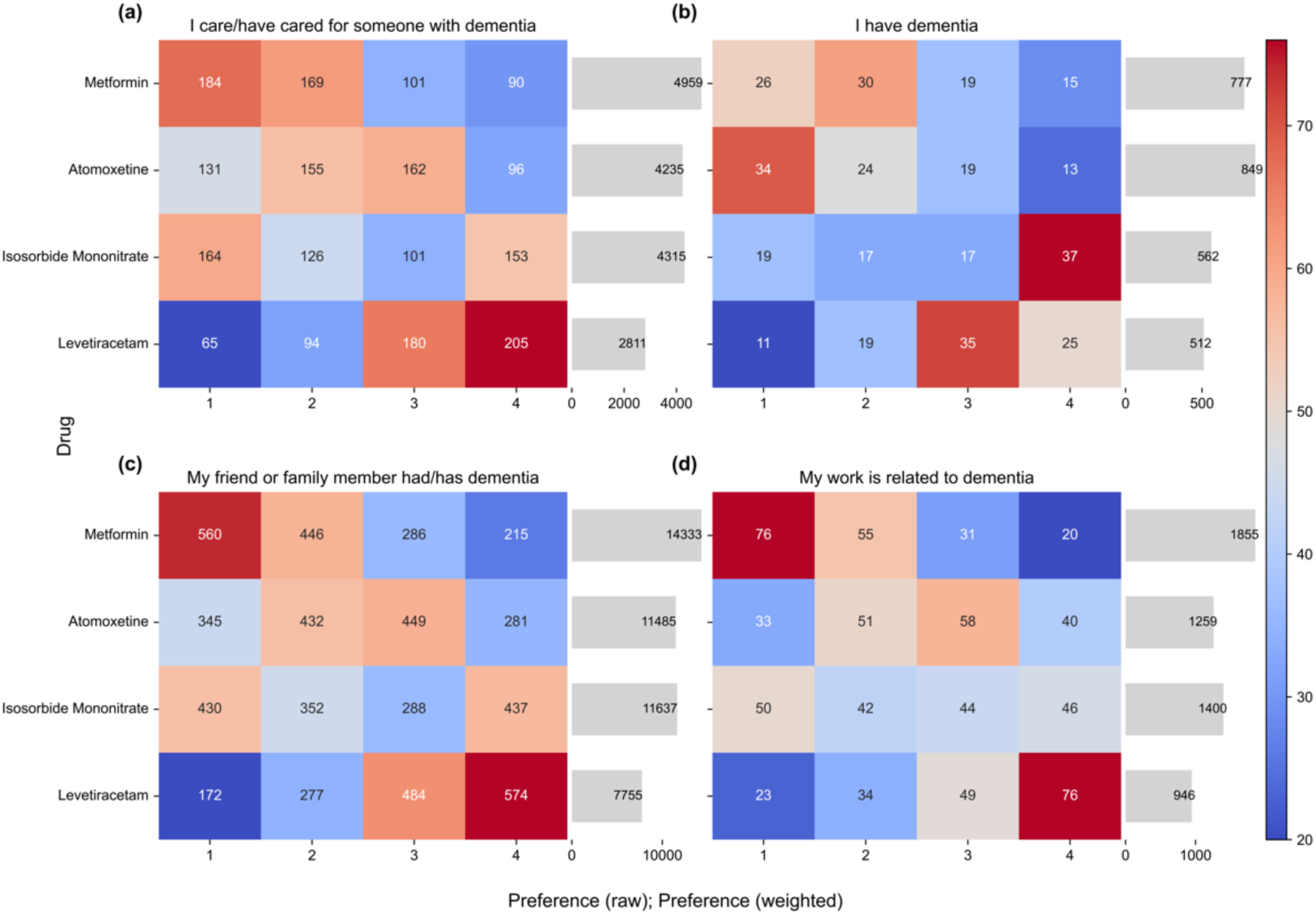
Preferences across participants’ role. The heatmaps show the distribution of raw ranking frequencies for each drug. The bar charts show the corresponding total weighted preference scores. Results are presented separately for four participant groups: (a) caregivers, (b) individuals with dementia, (c) friends or family of people with dementia, and (d) professionals working in dementia-related fields.

Across the subgroups, two distinct patterns emerged. Participants who had caregiving experience (a), had a friend or family member with dementia (c), or worked in dementia-related fields (d) showed similar preference structures, where *Metformin* was consistently the most preferred drug, followed by *Isosorbide Mononitrate* and *Atomoxetine*, and *Levetiracetam* was the least favored. In contrast, participants living with dementia themselves (b) showed a different pattern, with *Atomoxetine* slightly surpassing *Metformin*, and the other two drugs receiving the lowest scores.

### 3.3. DCE results

#### 3.3.1. Overall results

We estimated a mixed logit model to examine public preferences across five treatment attributes (see Supplementary Table S3). Relative importance of each attribute was calculated based on the range of their estimated coefficients. Figure 5 displays the marginal utilities of the attribute levels.

**Figure 5.**
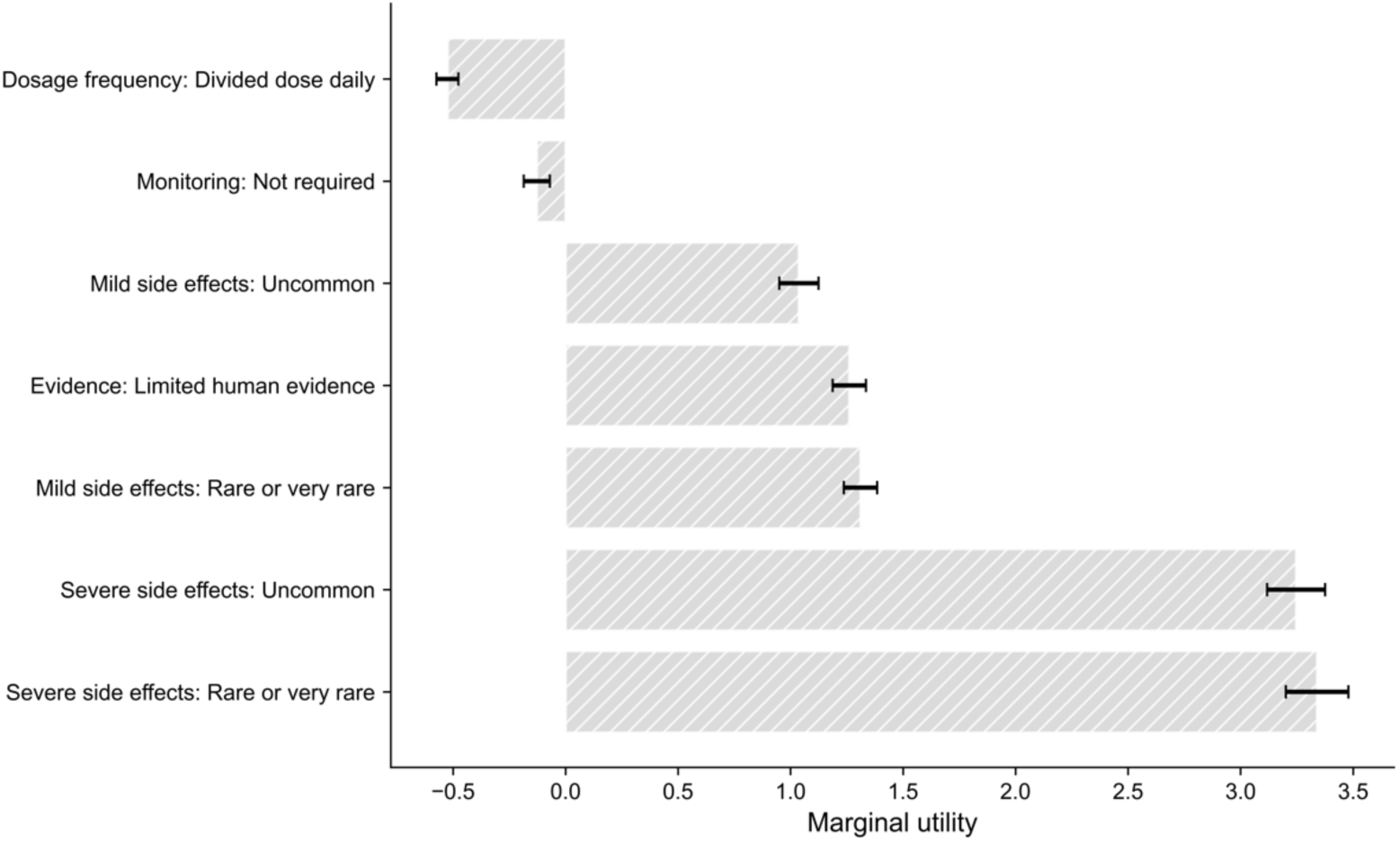
Marginal utilities of attribute levels. Error bars represent the estimated marginal utility for each attribute level relative to its reference level. Reference levels were: Dosage frequency: once daily; Monitoring: required; Evidence: pre-clinical animal trials; Mild side effects: common or very common; Severe side effects: common or very common. Error bars denote 95% CI.

All five attributes had a statistically significant effect on participants’ choice. Among these, severe side effects exhibited the highest impact on decision-making (relative importance: 50.88%). Both levels of severe side effect (*Uncommon and Rare or very rare*) showed strong positive marginal utilities (*β* = 3.247 and 3.340, respectively; both *p* < 0.001), indicating that respondents placed the greatest weight on avoiding severe adverse outcomes when evaluating medications. Similarly, mild side effects were found to be influential (relative importance: 19.97%), with significant preferences for medications associated with less frequent mild side effects (*β* = 1.037 for *Uncommon* and *β* = 1.311 for *Rare or very rare*, both *p* < 0.001), suggesting sensitivity to the probability of experiencing mild discomfort.

The third most important factor was the type of evidence (relative importance: 19.21%). Specifically, respondents preferred medications supported by stronger human evidence over those based solely on pre-clinical animal data (*β* = 1.261, *p* < 0.001).

In contrast, dosage frequency (relative importance: 8.0%) and monitoring requirements (relative importance: 1.95) had relatively small effects. Divided daily dosing (*β* = −0.525, *p* < 0.001) and not requiring monitoring (*β* = −0.128 , *p* = 0.029 ) were both associated with lower utility, suggesting a modest preference for simplicity and reduced burden.

Overall, these findings reveal a clear preference among respondents, with a strong emphasis on safety (especially avoiding severe side effects), followed by effectiveness (evidence credibility) and convenience (dosing and monitoring).

#### 3.3.2. Subgroup analyses

##### Age and gender

Figure 6 presents subgroup analyses stratified by age and gender, including four groups: males aged <65 years, males aged ≥65 years, females aged <65 years, and females aged ≥65 years.

**Figure 6.**
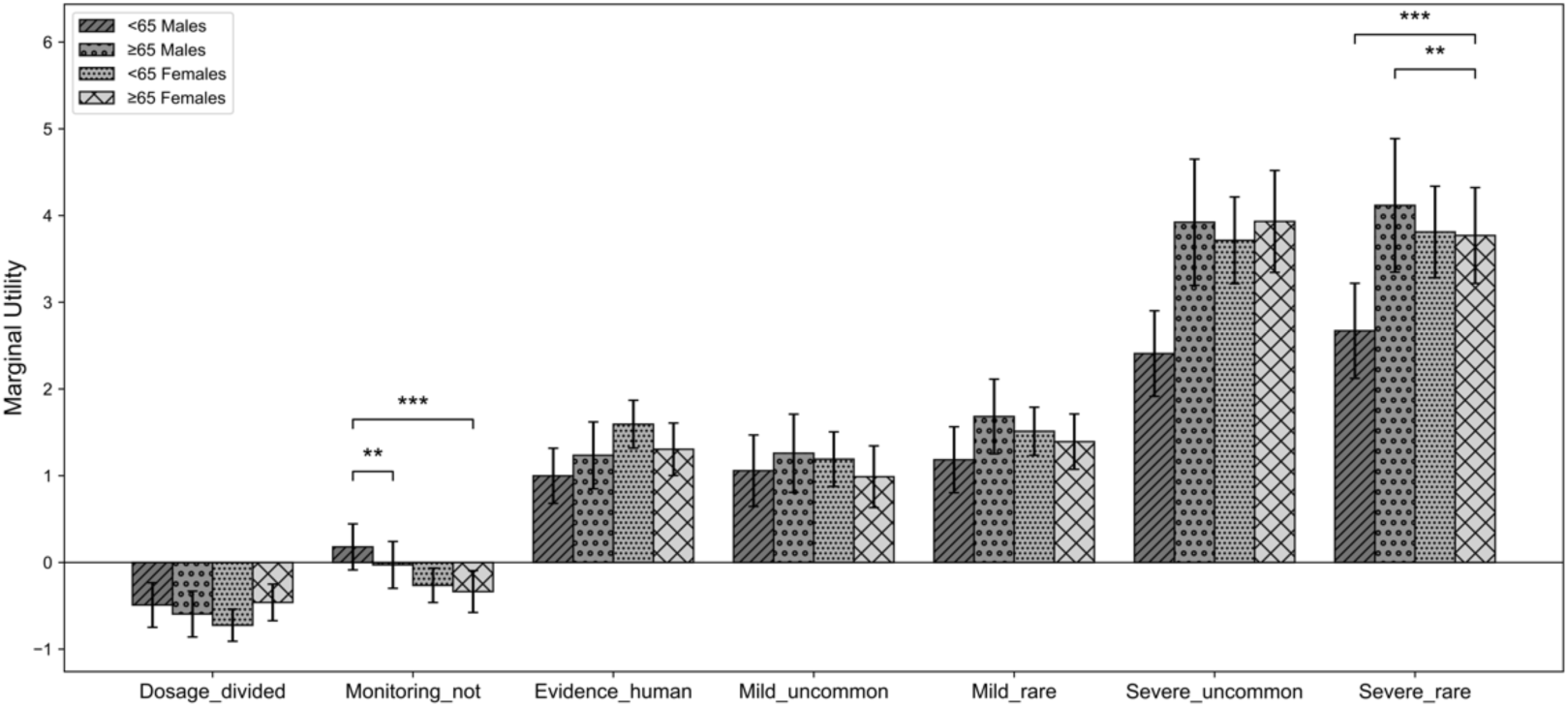
Subgroup marginal utilities with 95% CI. Bars represent four demographic groups (<65 males, ≥65 males, <65 females, ≥65 females). Significant pairwise differences are indicated by asterisks (∗ *p* < 0.05,∗∗ *p* < 0.01,∗∗∗ *p* < 0.001).

Across all groups, attributes related to side effects (both *uncommon* or *rare*) remained the strongest positive influence on treatment choice, followed by the strength of supporting evidence, whereas convenience-related attributes (dosage frequency and monitoring requirements) contributed less.

Notably, for the monitoring requirement, both male subgroups (<65 and ≥65 years old) did not exhibit statistically significant preferences. In contrast, both female subgroups demonstrated significant preferences for medications involving regular monitoring. The group comparison tests further revealed a significant difference between younger and older males. Specifically, younger males (<65 years old) showed a greater preference for drugs that do not require monitoring, whereas older males (≥65 years old) tended to prefer those that involve regular monitoring (*p* < 0.01). This finding suggests a potential age-related divide among males in terms of their acceptance of ongoing health monitoring. In addition, we also observed a significant difference between younger males and older females (*p* < 0.001). Younger males (<65 years old) appeared more inclined to avoid frequent monitoring, while older females (≥65 years old) were more accepting of regular health monitoring.

In addition, the magnitude of preferences for severe side effects varied by age and gender. Younger males (<65 years old) and older females (≥65 years old) differed significantly in their preferences, with older females showing a stronger aversion to severe side effects, suggesting a greater emphasis on drug safety among older women ( *p* < 0.001 ). Additionally, we observed a significant difference between older males and older females (≥65 years old). The results showed that men in the older age group place even more weight on avoiding severe side effects compared to their female counterparts (*p* < 0.01).

##### Dementia-related experience

Figure 7 further illustrates the estimated marginal utilities and 95% CI for groups with different dementia experience: I have dementia, I care/have cared for someone with dementia, my friend or family member had/has dementia, and my work is related to dementia.

**Figure 7.**
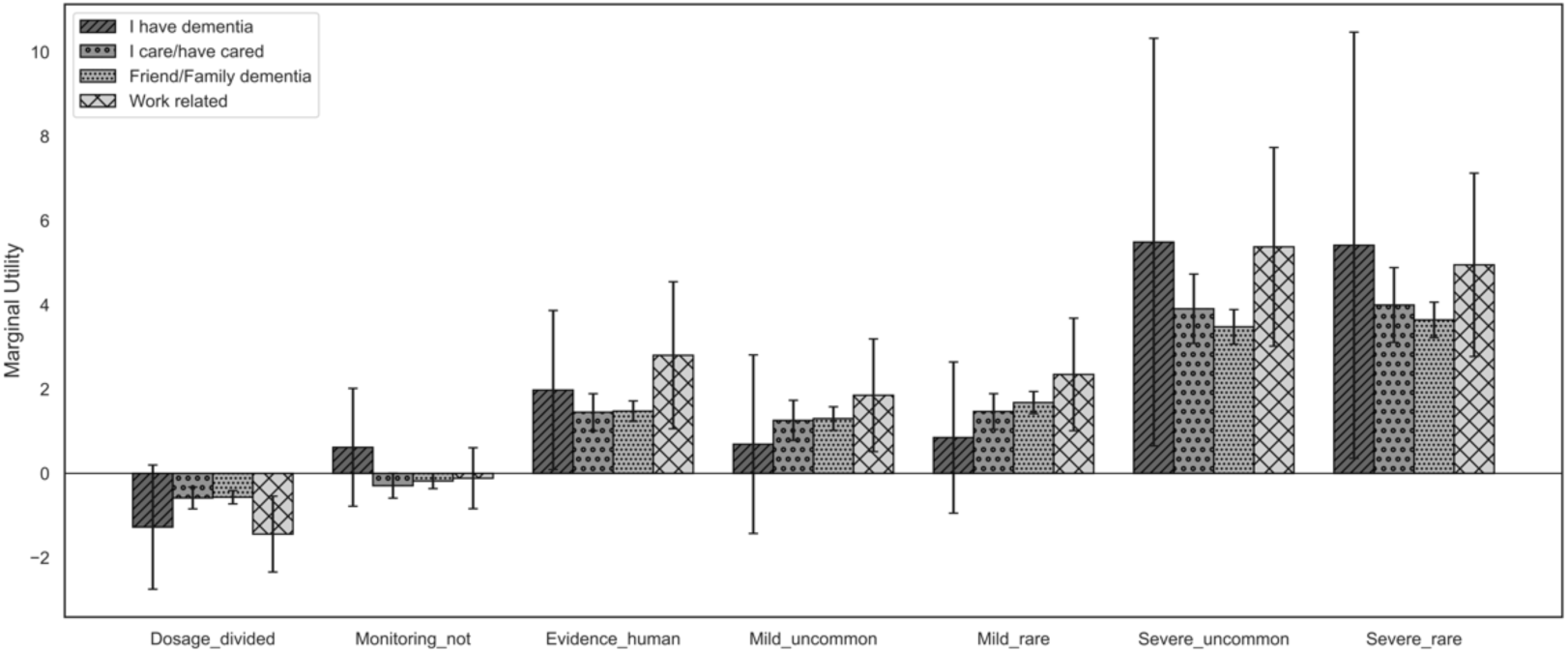
Subgroup marginal utilities with 95% CI. Bars represent four groups with different dementia experience. Significant pairwise differences are indicated by asterisks (∗ *p* < 0.05,∗∗ *p* < 0.01,∗∗∗ *p* < 0.001).

The overall preference patterns across dementia-related experience were broadly consistent with those observed in the full sample. Side effects remained the strongest determinants, followed by evidence strength, whereas convenience-related attributes contributed minimally.

Although no statistically significant differences were detected between subgroups, a notable finding was observed for the group living with dementia. For these participants, only severe side effects and evidence strength had a statistically significant positive influence on treatment preferences, while mild side effects and convenience-related attributes did not significantly affect their decision-making.

## 4. Discussion

### 4.1. Summary

We have demonstrated it is possible to use a web-based portal to engage with patients and the public at scale to help design a research project in dementia. We were also able to understand views on our research approach, obtain rankings for four compounds for repurposing in Alzheimer’s Disease and, crucially, understand factors that are most important in making these decisions. We found that people were enthusiastic about the idea of a platform trial in AD, with 79.6% of participants reporting very positive or somewhat positive attitudes. Among the four candidate drugs, they favoured *Metformin*, followed by *Atomoxetine* and *Isosorbide Mononitrate* and were less enthusiastic about *Levetiracetam*. The main driver of decision making was avoiding serious side effects, followed by mild side effects and strong supporting evidence of efficacy, and convenience (for example single daily dosing or not needing blood test monitoring). We were able to examine these results by respondent characteristics (sex, age, experience of dementia). The findings from these subgroup analyses were generally consistent with those of the main analysis.

### 4.2. Strengths and limitations

The major strength of our study is its large and diverse sample, eliciting the views of more than 3,000 people from all over the world. Our digital platform complements traditional small scale PPIE addressing its common limitations in reach and representativeness. This approach enabled broader engagement by removing geographic and logistical barriers, offering multilingual access, and creating opportunities for large numbers of individuals to express their views. Moreover, scaling PPIE survey activity enabled us to go beyond simply capturing opinions, allowing meaningful inference about interaction effects and providing insights into the factors that make certain medications more or less attractive to take. Feedback on the survey experience also indicated that the vast majority of participants found it easy to complete, with only around 10% reporting any difficulty. This suggests that the survey was broadly accessible and that the co-design input from PPIE contributors helped ensure usability.

Another important strength lies in the cost-effectiveness of this approach. Once established, our platform allowed large-scale engagement at minimal additional cost. Participation was entirely voluntary and with no financial incentives offered, yet engagement remained high. This demonstrates strong public willingness to contribute to dementia research when the process is accessible and meaningful, reinforcing the value of web-based PPIE as a sustainable model.

Third, this study also provided valuable experience in the governance of this type of research, which required coordination of university ethics approval, sponsorship, and insurance for the website portal. Through the process, we also developed practical insights into effective recruitment strategies. While many initial approaches led to limited gains, adoption of the study by the NIHR Join Dementia Research (JDR) initiative led to a substantial increase in survey completions (https://www.joindementiaresearch.nihr.ac.uk/).^25^ This increase is illustrated in Supplementary Figure S1, which shows a marked rise in completions after the study went live on JDR on 26 June 2025.

Finally, more than half of the respondents agreed to be contacted again, establishing a unique cohort of nearly 2,000 individuals who can be directly reached in future PPIE activities. The POPPED portal is a resource available for future studies, at least four are already planning to use it.

Our study does have some limitations. The most obvious is the possibility of digital exclusion, which requires internet access and digital literacy. For those who are less comfortable with technology, including some older adults or people with limited access to digital devices, may therefore have been underrepresented. In our study, the digital exclusion may also partly account for the relatively low participation of people with dementia (n=90). Whilst this number is small, hearing the views of 90 people with dementia is important and it is also helpful to be able to identify their views separately to carers or others. It is also likely other groups were excluded, for example those who do not speak either English or Mandarin Chinese. Expanding language accessibility in future studies would help to capture more diverse perspectives.

In addition, the small size of some subgroups, particularly individuals living with dementia, may limit the statistical power of subgroup analyses and may have caused potential sampling bias. Finally, while the survey questions were carefully designed, they cannot fully capture the complexity of participants’ reasoning and may have influenced responses through the framing of choices. Future studies should complement survey findings with qualitative approaches to gain deeper insights.

### 4.3. Future study

Our study represents a complementary approach aimed at expanding the reach and inclusivity of public input. We hope others will use the POPPED portal and it will become an important and useful tool for PPIE in dementia research. The project has also raised new questions for future investigation. For instance, in our study, findings from the small qualitative group were broadly consistent with those from the large-scale PPIE survey. However, it remains unclear how researchers should interpret results when these approaches diverge. For example, if there were disagreements about which drugs to include. How should we weigh a rich qualitative discussion with a small number of people against an impersonal survey with a much larger sample size? Determining how to balance these complementary sources of evidence warrants further exploration.

In addition, we were pleased to be able to demonstrate that we could use the platform to obtain additional insights on factors that influenced drug preference, through the use of DCEs. The portal highlights its potential to address many more questions in the future. Lastly, future work should focus on understanding and minimising digital exclusion, while acknowledging that this portal is designed to complement others approaches to PPIE.

In summary, we present what we believe to be the largest PPIE exercise conducted to support a single study in Alzheimer’s disease. It will provide insights which will directly inform a forthcoming major trial platform and is a sustainable infrastructure which could be used by many future studies, improving their quality and ultimately care and treatment.

## Data Availability

All data produced in the present study are available upon reasonable request to the authors

## Acknowledgements

We thank the AD-SMART PPIE group for their support. We thank Dr Mark Toshner and Joe Newman for their advice in the governance around web based PPIE projects. A full list of members is provided in the Supplementary Material S.3.

## Consent Statement

All participants in the survey provide electronic consent prior to participation.

