## Supplementary Materials for "PPIE in clinical trials at scale: analysis of the first 3,250 responses on the POrtal for Patient and Public Engagement in Dementia (POPPED)"

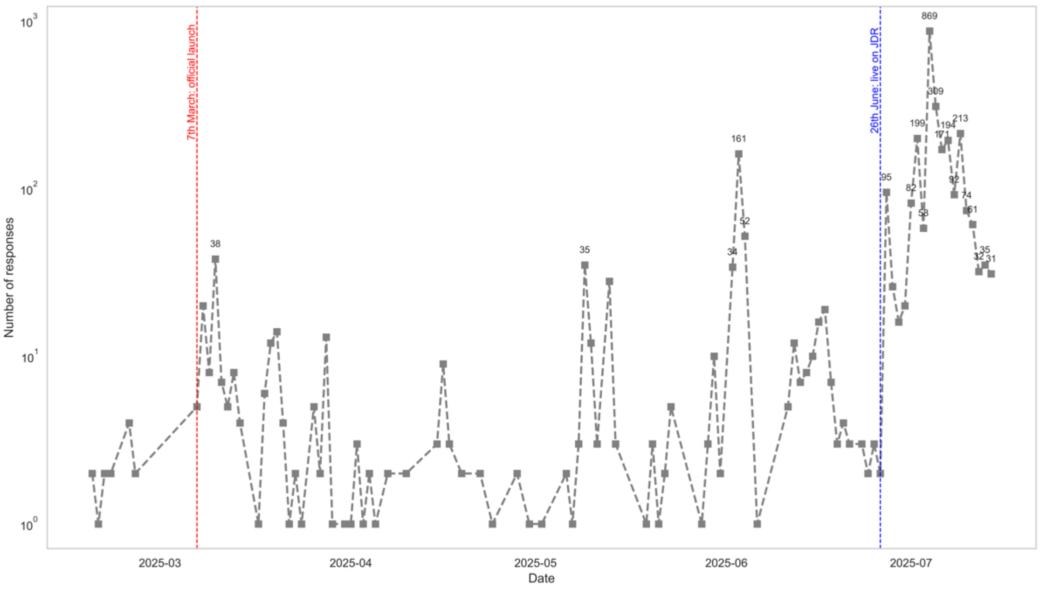


**FIGURE S1 Survey responses over time.** Daily number of survey completions (logarithmic scale) from March to July 2025. The red dashed line marks the launch of the survey website, and the blue dashed line marks the study's appearance on the Join Dementia Research (JDR) platform on 26 June 2025.

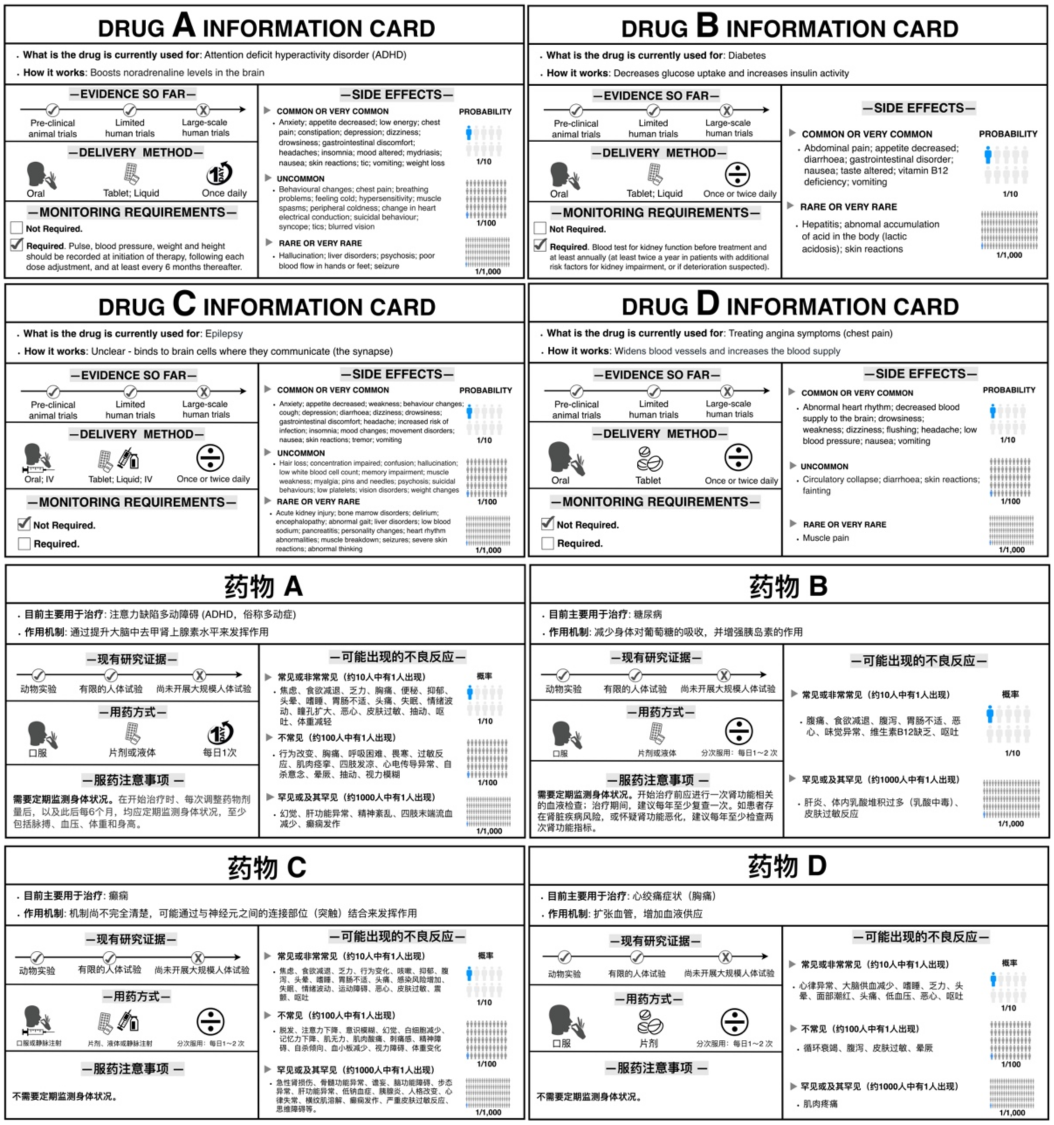


**FIGURE S2 Detailed drug cards in English and Mandarin Chinese.**

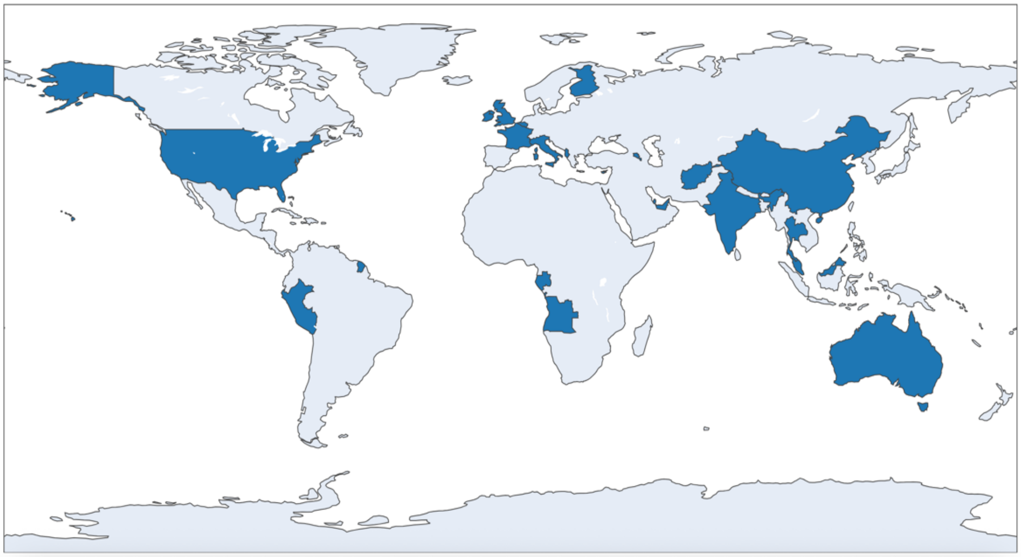

**FIGURE S3 Number of responses by country or region.**

**TABLE S1 Attributes and levels in the DCE element.**

| Attributes | Definition | Levels of attributes | Description |
|---|---|---|---|
| Dosage frequency | The amount of medication taken at a specific time. | Once daily | Only need to be taken once a day. |
| | | Divided dose daily | The total dose should be divided into several doses per day. |
| Monitoring requirements | Whether people need medical testing and monitoring before and after taking the drug. | Required | People are required to undergo medical tests and monitoring, such as liver function tests, kidney function tests, and blood tests, before and after using the medication. |
| | | Not required | People are not required to undergo additional medical tests before and after using the medication. |

| Attributes | Definition | Levels of attributes | Description |
|---|---|---|---|
| Evidence type | The type or stages of research conducted, ranging from pre-clinical studies to limited human evidence. | Pre-clinical animal evidence | Evidence comes solely from animal models or in vitro studies, with no human trials yet. |
| | | Limited human evidence | Small-scale or early-phase human studies suggest potential benefits but lack robust results. |
| Probability of mild short-term side effects | These include common side effects that are generally mild and may resolve on their own within a short period. These may include: dizziness, nausea, diarrhea, and headaches. | Very common and common | 1 out of 10 people |
| | | Uncommon | 1 out of 100 people |
| | | Rare or very rare | 1 out of 1,000 people |
| Probability of severe side effects | Severe side effects that may necessitate medical intervention or hospitalization. These may include severe palpitations, nephropathy, lactic acidosis, reversible renal impairment, respiratory disorders, fainting, severe allergic reactions, cardiac arrhythmias, and severe hepatic dysfunction. | Very common and common | 1 out of 10 people |
| | | Uncommon | 1 out of 100 people |
| | | Rare or very rare | 1 out of 1,000 people |

**TABLE S2 Sixteen choice tasks selected in the experimental design.**

| Block | Dosage frequency | Monitoring requirement | Efficacy evidence | Mild side effects | Severe side effects |
|---|---|---|---|---|---|
| 1 | Divided dose daily | Required | Pre-clinical animal trials | Rare or very rare | Uncommon |
| | Once daily | Not required | Limited human trials | Common or very common | Rare or very rare |

| Block | Dosage frequency | Monitoring requirement | Efficacy evidence | Mild side effects | Severe side effects |
|---|---|---|---|---|---|
| | Divided dose daily | Not required | Pre-clinical animal trials | Rare or very rare | Rare or very rare |
| | Once daily | Required | Limited human trials | Uncommon | Common or very common |
| | Once daily | Required | Pre-clinical animal trials | Common or very common | Uncommon |
| | Divided dose daily | Not required | Limited human trials | Uncommon | Common or very common |
| | Divided dose daily | Required | Pre-clinical animal trials | Rare or very rare | Uncommon |
| | Once daily | Not required | Limited human trials | Common or very common | Rare or very rare |
| 2 | Once daily | Required | Limited human trials | Rare or very rare | Common or very common |
| | Divided dose daily | Not required | Pre-clinical animal trials | Uncommon | Uncommon |
| | Once daily | Required | Limited human trials | Rare or very rare | Uncommon |
| | Divided dose daily | Not required | Pre-clinical animal trials | Uncommon | Common or very common |
| | Divided dose daily | Required | Limited human trials | Common or very common | Uncommon |
| | Once daily | Not required | Pre-clinical animal trials | Rare or very rare | Rare or very rare |
| | Once daily | Required | Limited human trials | Rare or very rare | Common or very common |
| | Divided dose daily | Not required | Pre-clinical animal trials | Uncommon | Uncommon |
| 3 | Once daily | Not required | Pre-clinical animal trials | Uncommon | Uncommon |
| | Divided dose daily | Required | Limited human trials | Common or very common | Common or very common |
| | Divided dose daily | Not required | Limited human trials | Common or very common | Rare or very rare |

| Block | Dosage frequency | Monitoring requirement | Efficacy evidence | Mild side effects | Severe side effects |
|---|---|---|---|---|---|
| | Once daily | Required | Pre-clinical animal trials | Uncommon | Common or very common |
| | Once daily | Required | Pre-clinical animal trials | Common or very common | Rare or very rare |
| | Divided dose daily | Not required | Limited human trials | Rare or very rare | Uncommon |
| | Once daily | Not required | Pre-clinical animal trials | Uncommon | Uncommon |
| | Divided dose daily | Required | Limited human trials | Common or very common | Common or very common |
| 4 | Divided dose daily | Required | Pre-clinical animal trials | Rare or very rare | Rare or very rare |
| | Once daily | Not required | Limited human trials | Common or very common | Uncommon |
| | Divided dose daily | Required | Limited human trials | Uncommon | Rare or very rare |
| | Once daily | Not required | Pre-clinical animal trials | Rare or very rare | Common or very common |
| | Once daily | Required | Limited human trials | Uncommon | Rare or very rare |
| | Divided dose daily | Not required | Pre-clinical animal trials | Common or very common | Common or very common |
| | Divided dose daily | Required | Pre-clinical animal trials | Rare or very rare | Rare or very rare |
| | Once daily | Not required | Limited human trials | Common or very common | Uncommon |

Note: Based on drug characteristics and level, there were 72 hypothetical medications and 2,556 possible choice tasks between two hypothetical drugs. A D-efficient experimental design was created with STATA software to reduce the number of choice tasks to twelve. In addition, a repeated choice set was used to check the consistency of respondents' choices.

**TABLE S3 Mixed logit model result (full sample).**

| | |
|---|---|
| Mixed logit model | Number of obs = 23,056 |
| | LR chi2(7) = 1052.95 |
| Log likelihood = -5738.6682 | Prob > chi2 = 0.0000 |

| chosen | Coef. | Std. Err. | z | P>\|z\| | [95% Conf.Interval] | |
|---|---|---|---|---|---|---|
| **Mean** | | | | | | |
| dosage_divided | -0.525 | 0.049 | -10.670 | **0.000** | -0.622 | -0.429 |
| monitoring_not | -0.128 | 0.058 | -2.190 | **0.029** | -0.242 | -0.013 |
| evidence_human | 1.261 | 0.074 | 17.130 | **0.000** | 1.116 | 1.405 |
| mild_uncommon | 1.037 | 0.087 | 11.890 | **0.000** | 0.866 | 1.208 |
| mild_rare | 1.311 | 0.074 | 17.620 | **0.000** | 1.165 | 1.457 |
| severe_uncommon | 3.247 | 0.129 | 25.230 | **0.000** | 2.995 | 3.500 |
| severe_rare | 3.340 | 0.139 | 24.030 | **0.000** | 3.068 | 3.613 |
| **SD** | | | | | | |
| dosage_divided | 0.663 | 0.131 | 5.070 | **0.000** | 0.407 | 0.919 |
| monitoring_not | 1.828 | 0.097 | 18.840 | **0.000** | 1.637 | 2.018 |
| evidence_human | 1.641 | 0.122 | 13.460 | **0.000** | 1.402 | 1.880 |
| mild_uncommon | 1.803 | 0.283 | 6.380 | **0.000** | 1.249 | 2.357 |
| mild_rare | 0.409 | 0.199 | 2.050 | **0.040** | 0.019 | 0.800 |
| severe_uncommon | 0.202 | 0.284 | 0.710 | 0.476 | -0.355 | 0.759 |
| severe_rare | 0.766 | 0.320 | 2.390 | **0.017** | 0.139 | 1.393 |

Note: all categorical variables were dummy-coded.
Dosage: Once daily (baseline)
Monitoring: Required (baseline)
Evidence: Pre-clinical animal trials (baseline)
Mild side effects: Common or very common (baseline)
Severe side effects: Common or very common (baseline)
A positive coefficient indicates a higher preference compared to the baseline level.

### S.1 Study sample

The minimum sample size required for the DCE was determined using a standard parametric approach for choice probability estimation:

$$n \geq \frac{(1-p)}{rpa^2} \times \left(\Phi^{-1}\left(1 - \frac{\alpha}{2}\right)\right)^2$$

where $p$ is the expected true population probability (set to 0.5 for maximum variance), $r$ is the number of choice tasks completed per respondent (*3* in our design), $a$ is the acceptable margin of error around the true population probability (0.05), and $\Phi^{-1}$ is the inverse of the cumulative normal distribution function, and $\alpha$ is the significance level ($\alpha$ = 0.05). Therefore, a minimum of approximately 500 participants was required to maintain overall statistical power.

However, because we planned to conduct subgroup analyses by gender, age, respondent role and country, we needed to ensure that each subgroup would have an adequate sample size for meaningful comparisons. In particular, we focused on the largest stratification by age, which included eight groups (18-24, 24-34, 35-44, 45-64, 55-64, 65-74, 75-84, 85 and over). By targeting roughly 300 respondents per age group to ensure sufficient precision for meaningful comparisons. Allowing for an anticipated 20% exclusion, the overall sample size was set at around 2,900 participants to ensure sufficient statistical power for subgroup analyses robust comparisons across key demographic and contextual factors.

#### S.2 Model stability check

we conducted a post hoc model stability check to assess whether further data collection continued to improve the reliability of parameter estimates. Specifically, we re-estimated the model in stages, progressively increasing the number of respondents (e.g., 500, 1000, 1500, and 3000), and plotted the estimated coefficients for all attribute levels with 95% confidence intervals.

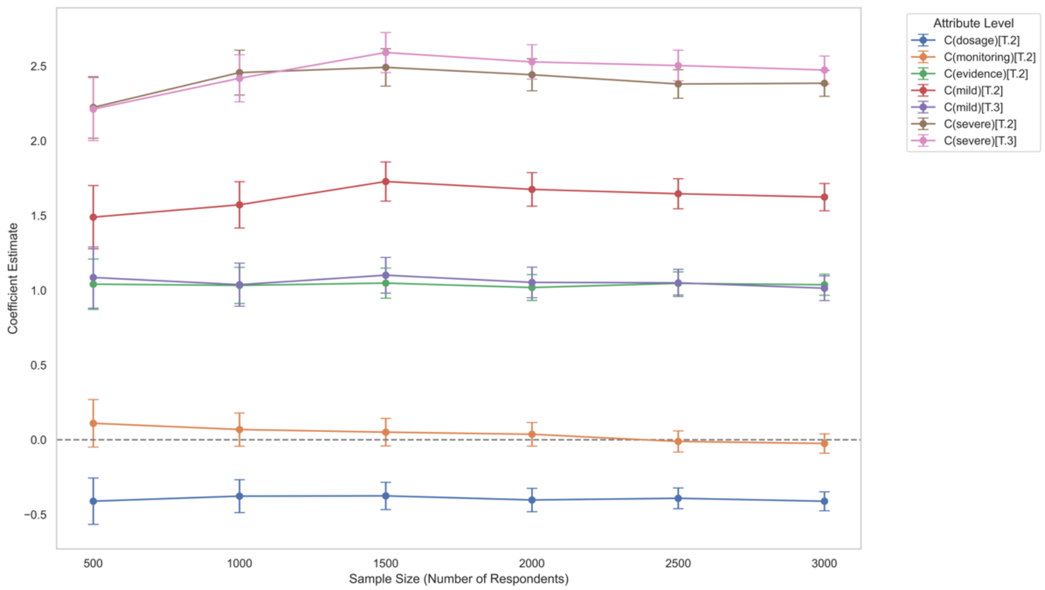


Figure S.2. Coefficient estimates by sample size

As shown in Figure S.2, most key coefficients stabilized after approximately 1000-1500 respondents, and confidence intervals narrowed with increasing sample size. This suggests that the data reached saturation well before the full sample size was collected, and additional responses had little impact on the model conclusions.

### S.3 Full list of AD-SMART contributors

#### AD-SMART Core Team

Professor Paresh Malhotra, Imperial College London -
Professor James Carpenter, MRC Clinical Trials Unit, University College London -
Professor Vanessa Raymont, University of Oxford -
Professor Ross Dunne, Greater Manchester Mental Health NHS Foundation Trust/University of Manchester -
Professor Suzanne Reeves, UCL -
Dr Benjamin R Underwood, University of Cambridge –
Linda Pointon, University of Cambridge -
Shabinah Ali, MRC Clinical Trials Unit -
Dr Sabahat Iqbal, Imperial College London -
Cristina Bonet-Olivares, Imperial College London -
Jo Whittle, MRC Clinical Trials Unit, University College London –
Laura Rizzo, Imperial College London -

#### Wider Team for Appendix

Fleur Hudson, MRC Clinical Trials Unit -
Isobel Landray, LSHTM -
Suzie Cro, Imperial College London -
Helen Brooks, UK DRI -
Dr Caroline S Clarke, UCL -
Lindsey Masters, MRC Clinical Trials Unit -
Sara Peres, MRC Clinical Trials Unit -
Carlos Diaz Montana, MRC Clinical Trials Unit -
Professor Max Parmar, MRC Clinical Trials Unit –
Professor Jeremy Chataway, MRC Clinical Trials Unit/UCL Queen Square -
Professor Siddharthan Chandran, UK DRI -
Professor Suvankar Pal, University of Edinburgh -
Professor Henrik Zetterberg, UK DRI -
Professor Dave Sharp, Imperial College London -
Professor Nick Fox -

Professor Liz Coulthard, University of Bristol -
Dr Amanda Heslegrave, UK DRI -
Professor Dame Louise Robinson, Newcastle University -
Professor Clive Ballard, University of Exeter -
Dr Timothy Rittman, University of Cambridge -
Dr Daniel Blackburn, University of Sheffield -
Professor Dag Aarsland, Kings College London -
Professor Alan Thomas, Newcastle University -
Professor Masud Husain, University of Oxford -
Professor James Rowe, University of Cambridge -
Professor Charles Marshall, Queen Mary University of London -
Dr Ivan Koychev, University of Oxford -
Dr Jay Amin, University of Southampton -
Dr Tom Russ, University of Edinburgh -
Dr Simon Bell, University of Sheffield -
Dr Jonathan Blackman, University of Bristol -
Dr Robert Barber, University of Newcastle -
Dr Tomas Welsh, University of Bristol -
Dr Zunera Khan, the Care Network
Professor John O'Brien -

**Drug prioritisation panel – independent members**
Professor Jeffrey Cummings -
Professor Fiona Ducotterd -
Professor Eric Karran -
Professor John O'Brien -
Professor Alastair Reith -
Professor Lon Schneider -
Professor Rik Vandenberghe -
Professor Gordon Wilcock -
Professor Cath Mummery -

**Independent PPIE members:**
Ajay Dave
Eric Deeson

Brian Human
Jakki Levene
Pamela Marie Lumbroso
Remy Olasoji
Heather Richardson
Ian Richardson
Martin Robertson
David Ross
Charlotte Scrimgeour
Sheila Wonnacott
David Winskill
Winnie Henry

**Charity Representatives**

Dr Zunera Khan, the Care Network
Amy Kordiak, Alzheimer's Society

Emma Wolverson, Dementia UK

Anna Goodman, ARUK

Dr Leah Mursaleen

Dr Sheona Scales
Dr Josie Waters
